# The impact of gastric acid suppression on the developing intestinal microbiome of a child

**DOI:** 10.1101/2021.12.21.21268064

**Authors:** Spurr Andrew, Zhu Wei, S. W. Wong Wendy, Diez Bernadette, Leibowitz Ian, Louis-Jacques Otto, Vilboux Thierry, Niederhuber John, L Levy Shira, K Hourigan Suchitra

## Abstract

Gastric acid suppressing medications have been associated with an increased risk of Clostridioides difficile (C. difficile) infection, hypothesized to be due to underlying intestinal microbiome changes. Our goal is to characterize these changes in children as their microbiome is undergoing critical development. Our study included 5 children (< 3 years old) who were started on clinically indicated gastric acid suppression and 15 healthy age-matched controls. Stool samples were collected before and after 2 months of treatment. We analyzed the microbiome using 16S rRNA sequencing. Quantitative-PCR was used to detect C. difficile toxins. Subjects and controls had similar alpha and beta diversity. We found no significant change in alpha or beta diversity of subjects after treatment. C. difficile toxins were not found and there was no increase in C. difficile carriage after treatment. A significant increase in Lactobacillus was found after treatment, which has been associated with C. difficile in adults.

## Introduction

The human intestinal microbiome contains at least 10^14^ bacteria (1,2), that exist in symbiosis with their host. This community of microorganisms performs vital functions for their host including development and maturation of the immune and metabolic system (3,4). Conversely, imbalance, or dysbiosis, of the microbiome has been associated with a wide range of diseases in humans, from gastrointestinal conditions such as inflammatory bowel disease (IBD) (5,6) and irritable bowel syndrome (7), to metabolic conditions including obesity (8), atopic conditions such as asthma, food allergies and eczema (9,10) and even autism (11).

The microbiome develops rapidly in the first few years of life which shapes future digestive and immune health (12,13). Young children are often prescribed medication to suppress gastric acid, which include proton pump inhibitors (PPIs) and histamine H2 receptor antagonists (H2RAs), for gastroesophageal reflux and other conditions (14). We are learning more about the potential side effects of these medications including an increased risk of *C. difficile* infection (CDI)(15–18), small bowel bacterial overgrowth (19) and increased risk of allergic disease in children (20). We hypothesize that medications that suppress gastric acid cause a dysbiosis or imbalance in the microbiome of an individual that increases the risk of these side effects. Gastric acid suppression has been associated with microbiome changes to some degree in adult studies (21–24). Freedberg *et al*. demonstrated a change in taxa previously associated with CDI including an increase in Enterococcaceae and Streptococcaceae and a decrease in Clostridiales (23). Hojo *et al*. demonstrated a significant increase in *Lactobacillus* and *Streptococcus* with PPI treatment (24). However, gut microbial changes with medication to suppress gastric acid has not been well examined in children, who may differ from adults due to a dynamic developing microbiome in the first few years of life compared to a relatively stable microbiome in adults. To the best of our knowledge the only study of the effects of gastric acid suppression on the gut microbiome of pediatric subjects was published by Castellani *et al*. in 2017 (25). Their study of 12 infants concluded that PPI therapy had only minor effects on the fecal microbiome that were no longer seen after discontinuation of therapy; however there was no control group of healthy untreated infants (25).

Understanding the effect of routinely used gastric acid suppression medications on the developing microbiome of the child will give insight into possible mechanisms for reported side effects and may also provide data to guide appropriate use of these commonly used medications in childhood. This pilot study aimed to examine the longitudinal fecal microbiome of young children prior to starting medication to suppress gastric acid and during therapy compared to healthy age-matched controls not receiving medication. In addition, we aimed to assess the presence of toxigenic *C. difficile* with treatment.

## Methods

Under the IRB approved protocol (#16-2215) at the Inova Health System, we enrolled children under the age of 3 years who were prescribed clinically indicated gastric acid suppression medication for gastroesophageal reflux symptoms by their treating physician. Parents provided written consent for participation in the study. Exclusion criteria included: antibiotic use in the preceding three months, use of the same class of medication to suppress gastric acid in the preceding three months, transitioning from a PPI to H2RA in clinic, and a consenting parent younger than 18 years of age. Demographic and clinical data were collected during the clinic visit at which they were first prescribed the medication. Stool from the subject was collected by placement on a fecal occult blood card, a method previously validated, at two time points (26). The first sample was collected prior to initiation of gastric acid suppression medication and second sample was collected two months later. Samples were mailed from home in FDA approved mailers. Fecal occult blood cards were stored at -80°C until analysis.

Age- and gender-matched control subjects who were considered healthy and were not on medication to suppress gastric acid were also included from a longitudinal cohort study within the Inova Health System (IRB protocol # 15-1804). Stool was collected in the same manner at one time point.

DNA was extracted from the fecal occult blood cards using the Qiagen EZ1 Advanced with the EZ1 Tissue Kit and the Bacterial DNA Extraction protocol card. Samples were cleaned and concentrated using the DNeasy PowerClean Cleanup kit (Qiagen). Sequencing libraries were prepared using a Nextera XT kit (Illumina, San Diego, CA) using a modified Illumina 16S Metagenomics Sequencing Library Preparation protocol for analysis of hypervariable region V4. Each sample was sequenced on the Illumina MiSeq with paired end reads of 301bp. Sequencing of negative controls of lysis buffer and positive controls of Staphylococcus aureus (Strain NCTC 8532, ATCC, VA) and Escherichia coli (Strain NCTC 9001, ATCC, VA) were included.

De-multiplexed sequences were filtered, trimmed, and clustered into operational taxonomic units (OTUs) using the *R* package *DADA2* (version 1.8). While this pipeline is available within the QIIME2 framework, it was used separately to allow for more customization of parameters. The sequence table and OTU table generated by *DADA2* were imported into QIIME2 (version 2018.2), where a phylogenetic tree was generated using *FastTree* and taxonomic identification was performed using the Greengenes 16S rRNA database (version gg 13_8 99% OTUs). The results were imported into the *R* package *phyloseq* (version 1.23.1) to perform alpha diversity (Shannon and Simpson) and beta diversity (weighted unifrac) analyses. Differential abundances were calculated using the *R* package *DESeq2* (version 1.18.1). Graphs and plots were generated using the *R* package *ggplot2* (version 2.2.1). The sequence data are being deposited in NCBI Sequence Read Archive (SRA).

Quantitative-PCR was used to detect 16S rRNA, *C. difficile* toxin A (TcdA), and *C. difficile* toxin B (TcdB) genes of *C. difficile* as previously described (27).

## Results

Five children were enrolled in the pilot study, 3 of whom were started on H2RAs and the remaining 2 on PPIs (Table 1). Two stool samples from each subject (“Before” treatment and “After” treatment) were included in the analysis. Fifteen age-matched controls were also included who were not on medication to suppress gastric acid (Table 2).

**Table 1:** Subject demographic and clinical information. Subject 4 was treated with antibiotics for 2 days while enrolled in the study. Age quartiles based on age range of 1 to 33 months.

| Subject ID | Gender | Age at first sample | Medication Type | Antibiotics | Diet |
| --- | --- | --- | --- | --- | --- |
| 1 | F | 4<sup>th</sup> quartile | PPI | None | Cow's milk, almond milk, solid food |
| 4 | M | 1<sup>st</sup> quartile | H2RA | 2 days | Cow's milk formula |
| 8 | M | 2<sup>nd</sup> quartile | H2RA | None | Cow's milk formula |
| 10 | F | 3<sup>rd</sup> quartile | H2RA | None | Cow's milk, solid food |
| 12 | M | 2<sup>nd</sup> quartile | PPI | None | Cow's milk formula |

**Table 2:** Demographics of controls with their corresponding subjects. Age quartiles based on age range of 1 to 33 months.

| Subject ID | Subject Age | Control ID | Gender | Age (months) |
| --- | --- | --- | --- | --- |
| 1 | 4<sup>th</sup> quartile | 001B_1 | FEMALE | 4<sup>th</sup> quartile |
|  |  | 001B_2 | FEMALE | 4<sup>th</sup> quartile |
|  |  | 001B_3 | FEMALE | 4<sup>th</sup> quartile |
| 4 | 1<sup>st</sup> quartile | 004B_1 | MALE | 3<sup>rd</sup> quartile |
|  |  | 004B_2 | MALE | 3<sup>rd</sup> quartile |
|  |  | 004B_3 | MALE | 3<sup>rd</sup> quartile |
| 8 | 2<sup>nd</sup> quartile | 008B_1 | MALE | 3<sup>rd</sup> quartile |
|  |  | 008B_2 | MALE | 3<sup>rd</sup> quartile |
|  |  | 008B_3 | MALE | 3<sup>rd</sup> quartile |
| 10 | 3<sup>rd</sup> quartile | 010B_1 | FEMALE | 3<sup>rd</sup> quartile |
|  |  | 010B_2 | FEMALE | 3<sup>rd</sup> quartile |
|  |  | 010B_3 | FEMALE | 4<sup>th</sup> quartile |
| 12 | 2<sup>nd</sup> quartile | 012B_1 | MALE | 3<sup>rd</sup> quartile |
|  |  | 012B_2 | MALE | 3<sup>rd</sup> quartile |
|  |  | 012B_3 | MALE | 3<sup>rd</sup> quartile |

### Alpha and Beta Diversity

Alpha diversity was calculated using the Shannon Index. There was no significant change in alpha-diversity before and after gastric acid suppression (*p* = 0.51, Figure 1). Beta-diversity was determined using PCoA unifrac (weighted). Samples from the same individual before and after treatment clustered well together. Samples also clustered well by gender. Samples did not cluster by type of gastric acid suppression medication, nor by time point (before vs. after taking medication) (Figure 1).

**Figure 1:**
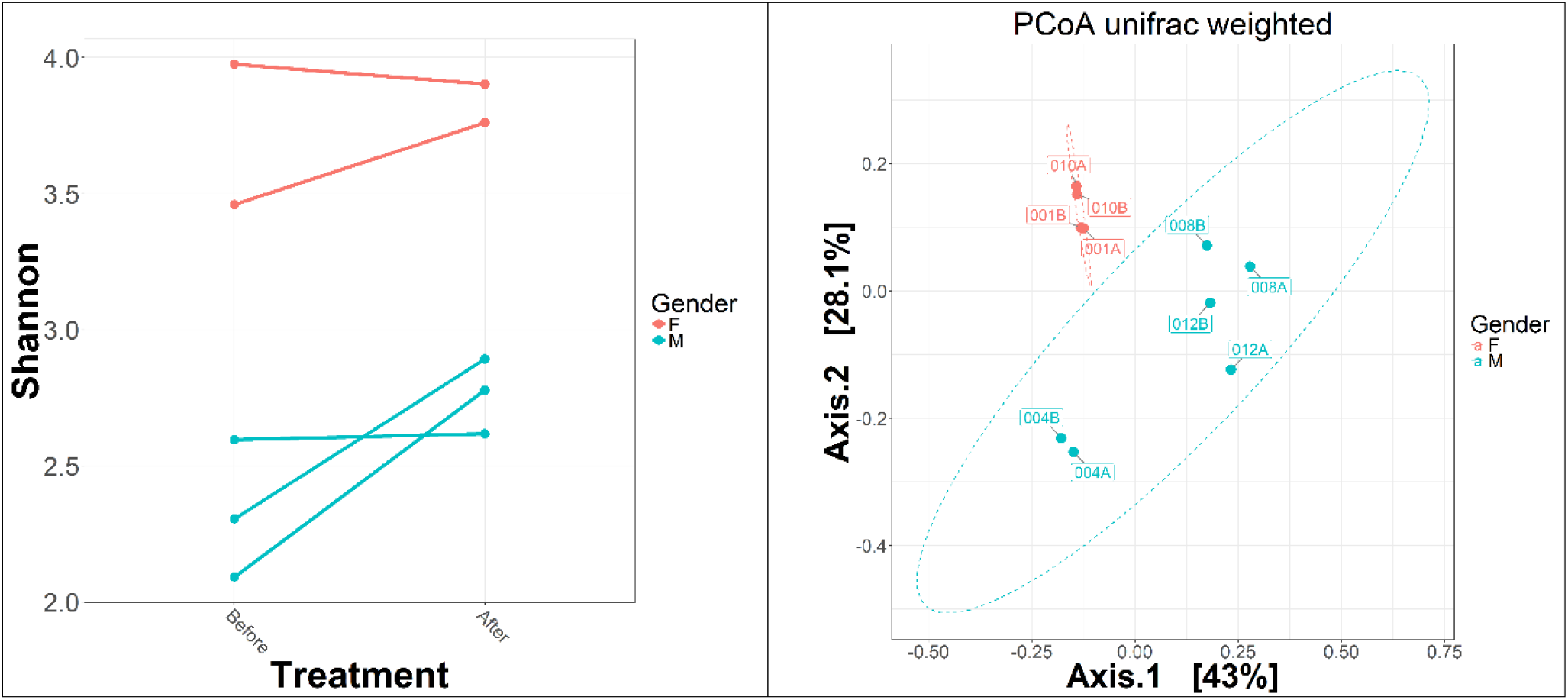
Shannon diversity (left) and beta diversity as measured by weighted unifrac (right). There was no significant change in Shannon diversity after gastric acid suppression. Samples clustered well by gender, however they did not cluster by type of medication, nor by time point.

### Relative Abundance of Taxa

Bacterial abundance before and after gastric acid suppression were calculated for each taxon (Table 3). Of note, the phylum Proteobacteria and the genus *Neisseria* significantly decreased after medication (*p* = 0.018, *p* = 0.028, respectively) and *Lactobacillus* significantly increased after medication (*p* = 0.002). Females had a significant decrease in *Enterococcus* (*p* = 0.024). Males had a significant decrease in *Enterobacter* (*p* = 0.038). Subjects taking H2RAs had a significant decrease in both *Haemophilus* and *Neisseria*. While only a select few taxa were significantly different when pooling different subjects together, there were more dramatic changes in taxa on an individual level after medication (Figure 2).

**Table 3:**
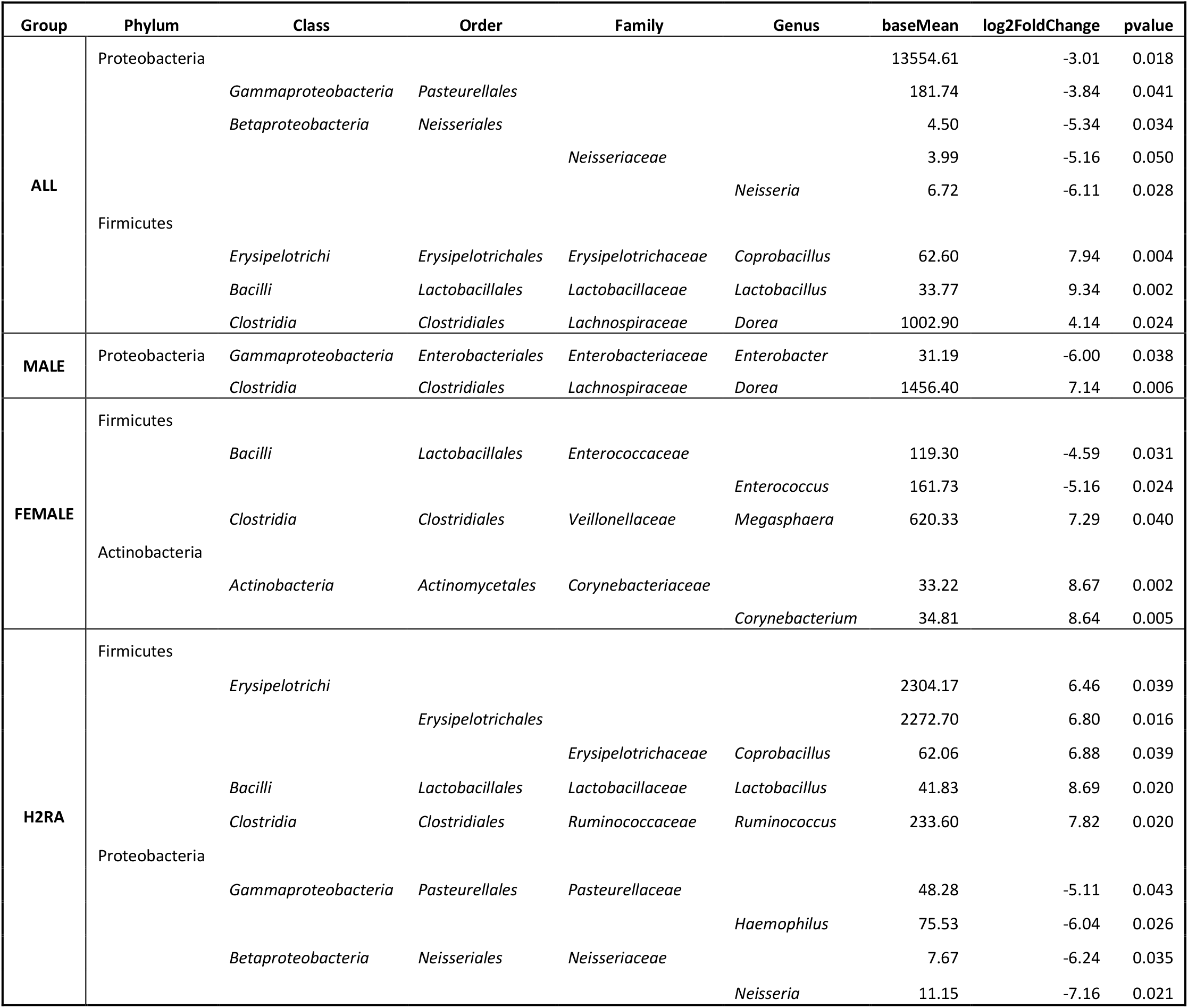
Significant changes in differential abundances from before and after medication use. Any taxon with a p-value had a significant change in abundance after medication. A negative log2 fold change indicates a decrease in abundance after medication and a positive log2 fold change indicates an increase in abundance after medication.

| Group | Phylum | Class | Order | Family | Genus | baseMean | log2FoldChange | pvalue |
| --- | --- | --- | --- | --- | --- | --- | --- | --- |
| ALL | Proteobacteria |  |  |  |  | 13554.61 | -3.01 | 0.018 |
|  |  | *Gammaproteobacteria* | *Pasteurellales* |  |  | 181.74 | -3.84 | 0.041 |
|  |  | *Betaproteobacteria* | *Neisseriales* |  |  | 4.50 | -5.34 | 0.034 |
|  |  |  |  | *Neisseriaceae* |  | 3.99 | -5.16 | 0.050 |
|  |  |  |  |  | *Neisseria* | 6.72 | -6.11 | 0.028 |
|  | Firmicutes |  |  |  |  |  |  |  |
|  |  | *Erysipelotrichi* | *Erysipelotrichales* | *Erysipelotrichaceae* | *Coprobacillus* | 62.60 | 7.94 | 0.004 |
|  |  | *Bacilli* | *Lactobacillales* | *Lactobacillaceae* | *Lactobacillus* | 33.77 | 9.34 | 0.002 |
|  |  | *Clostridia* | *Clostridiales* | *Lachnospiraceae* | *Dorea* | 1002.90 | 4.14 | 0.024 |
| MALE | Proteobacteria | *Gammaproteobacteria* | *Enterobacteriales* | *Enterobacteriaceae* | *Enterobacter* | 31.19 | -6.00 | 0.038 |
|  |  | *Clostridia* | *Clostridiales* | *Lachnospiraceae* | *Dorea* | 1456.40 | 7.14 | 0.006 |
| FEMALE | Firmicutes |  |  |  |  |  |  |  |
|  |  | *Bacilli* | *Lactobacillales* | *Enterococcaceae* |  | 119.30 | -4.59 | 0.031 |
|  |  |  |  |  | *Enterococcus* | 161.73 | -5.16 | 0.024 |
|  |  | *Clostridia* | *Clostridiales* | *Veillonellaceae* | *Megasphaera* | 620.33 | 7.29 | 0.040 |
|  | Actinobacteria |  |  |  |  |  |  |  |
|  |  | *Actinobacteria* | *Actinomycetales* | *Corynebacteriaceae* |  | 33.22 | 8.67 | 0.002 |
|  |  |  |  |  | *Corynebacterium* | 34.81 | 8.64 | 0.005 |
| H2RA | Firmicutes |  |  |  |  |  |  |  |
|  |  | *Erysipelotrichi* |  |  |  | 2304.17 | 6.46 | 0.039 |
|  |  |  | *Erysipelotrichales* |  |  | 2272.70 | 6.80 | 0.016 |
|  |  |  |  | *Erysipelotrichaceae* | *Coprobacillus* | 62.06 | 6.88 | 0.039 |
|  |  | *Bacilli* | *Lactobacillales* | *Lactobacillaceae* | *Lactobacillus* | 41.83 | 8.69 | 0.020 |
|  |  | *Clostridia* | *Clostridiales* | *Ruminococcaceae* | *Ruminococcus* | 233.60 | 7.82 | 0.020 |
|  | Proteobacteria |  |  |  |  |  |  |  |
|  |  | *Gammaproteobacteria* | *Pasteurellales* | *Pasteurellaceae* |  | 48.28 | -5.11 | 0.043 |
|  |  |  |  |  | *Haemophilus* | 75.53 | -6.04 | 0.026 |
|  |  | *Betaproteobacteria* | *Neisseriales* | *Neisseriaceae* |  | 7.67 | -6.24 | 0.035 |
|  |  |  |  |  | *Neisseria* | 11.15 | -7.16 | 0.021 |

**Figure 2:**
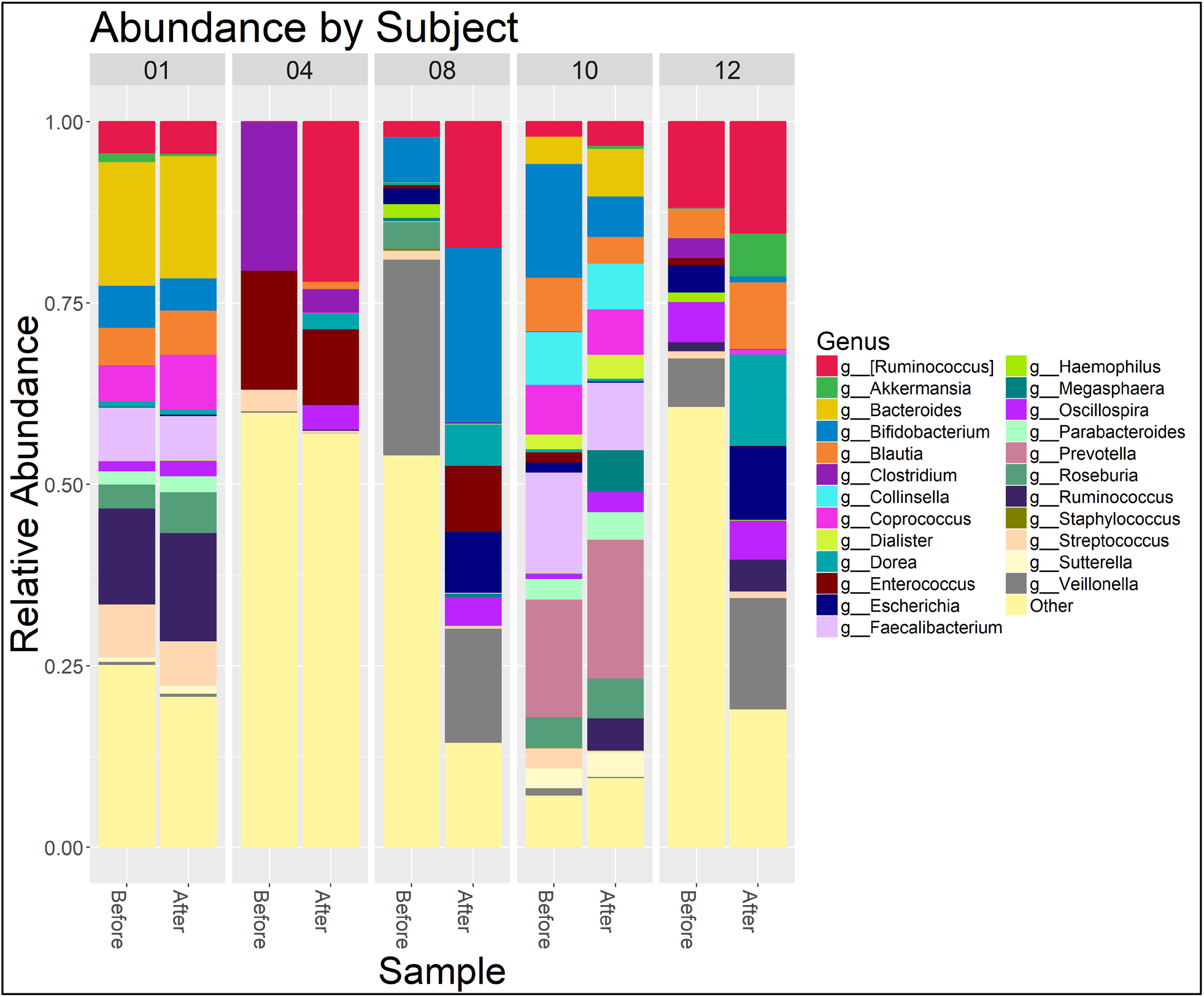
Relative abundance of the most abundant genera in each sample.

### *C. difficile* toxin

*C. difficile* toxin A (TcdA) and *C. difficile* toxin B (TcdB) were not found in any of the subjects. Nontoxigenic C. difficile was present only in one subject, both before and after PPI treatment.

### Control Comparisons

In order to account for age-and gender-related differences in microbial composition and diversity, we compared our subjects to three controls each of the same gender and similar age (Table 2). The controls had not been treated with antibiotics or medication to suppress gastric acid. They did not have clinically diagnosed gastroesophageal reflux disease.

#### Alpha diversity

The Shannon diversity, which takes into account not only the presence of certain OTUs, but the abundance as well, of the age- and gender-matched controls is similar to the diversity of the experimental samples (Figure 3, Table 4). The Shannon diversity of subjects and controls generally increases with age (Figure 4).

**Table 4:** P-values for mean Shannon Diversity of subjects vs. their age-matched controls.

| Subject | Mean Shannon Control Sample | Mean Shannon Study Sample | p value | Gender | Med |
| --- | --- | --- | --- | --- | --- |
| Subject 1 | 3.45 | 3.97 | 0.009 | F | PPI |
| Subject 4 | 2.92 | 2.61 | 0.003 | M | H2RA |
| Subject 8 | 3.34 | 2.45 | 0.176 | M | H2RA |
| Subject 10 | 3.66 | 3.68 | 0.903 | F | H2RA |
| Subject 12 | 3.24 | 2.62 | 0.252 | M | PPI |

**Figure 3:**
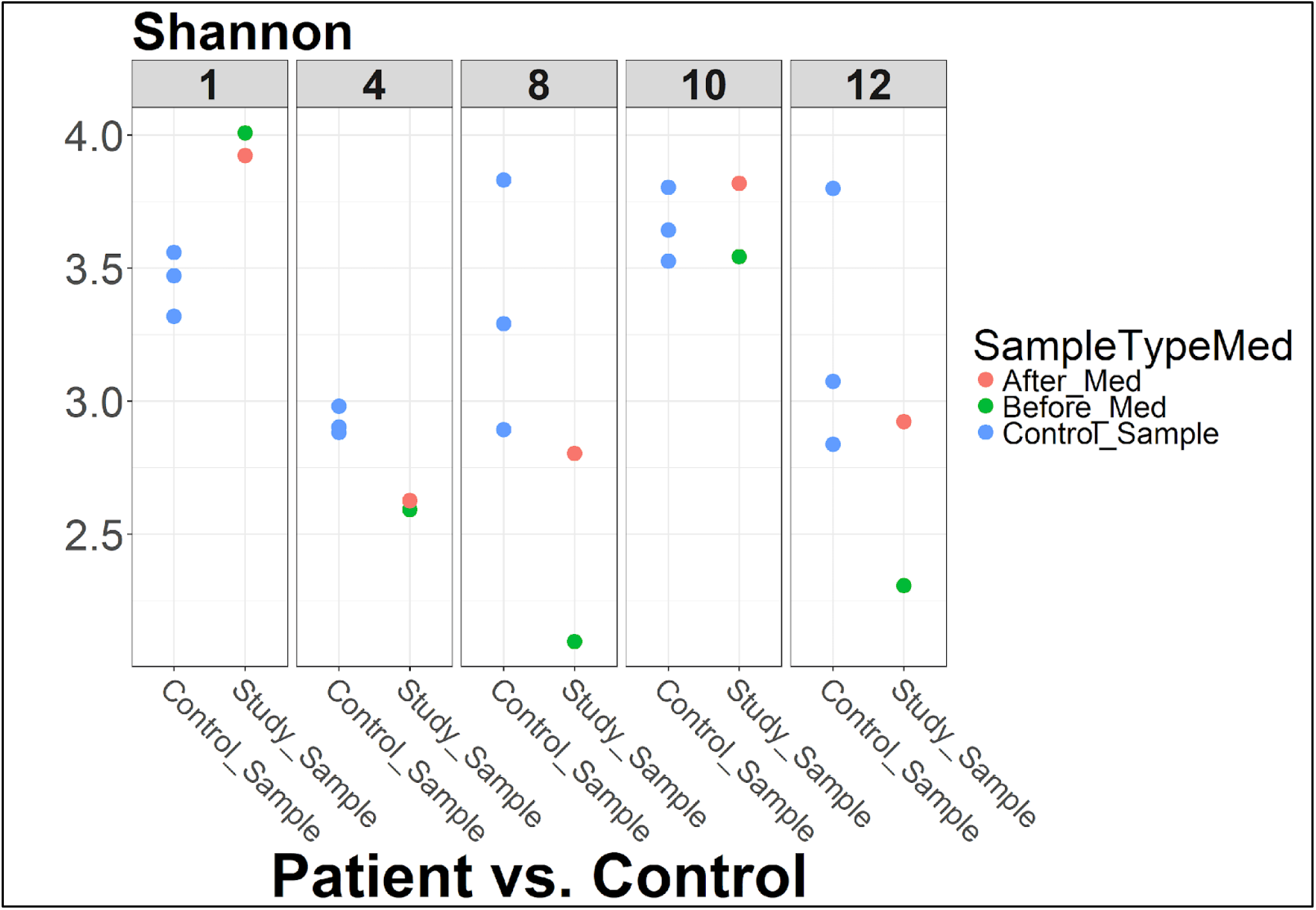
Comparison of alpha diversity to age- and gender-matched controls.

**Figure 4:**
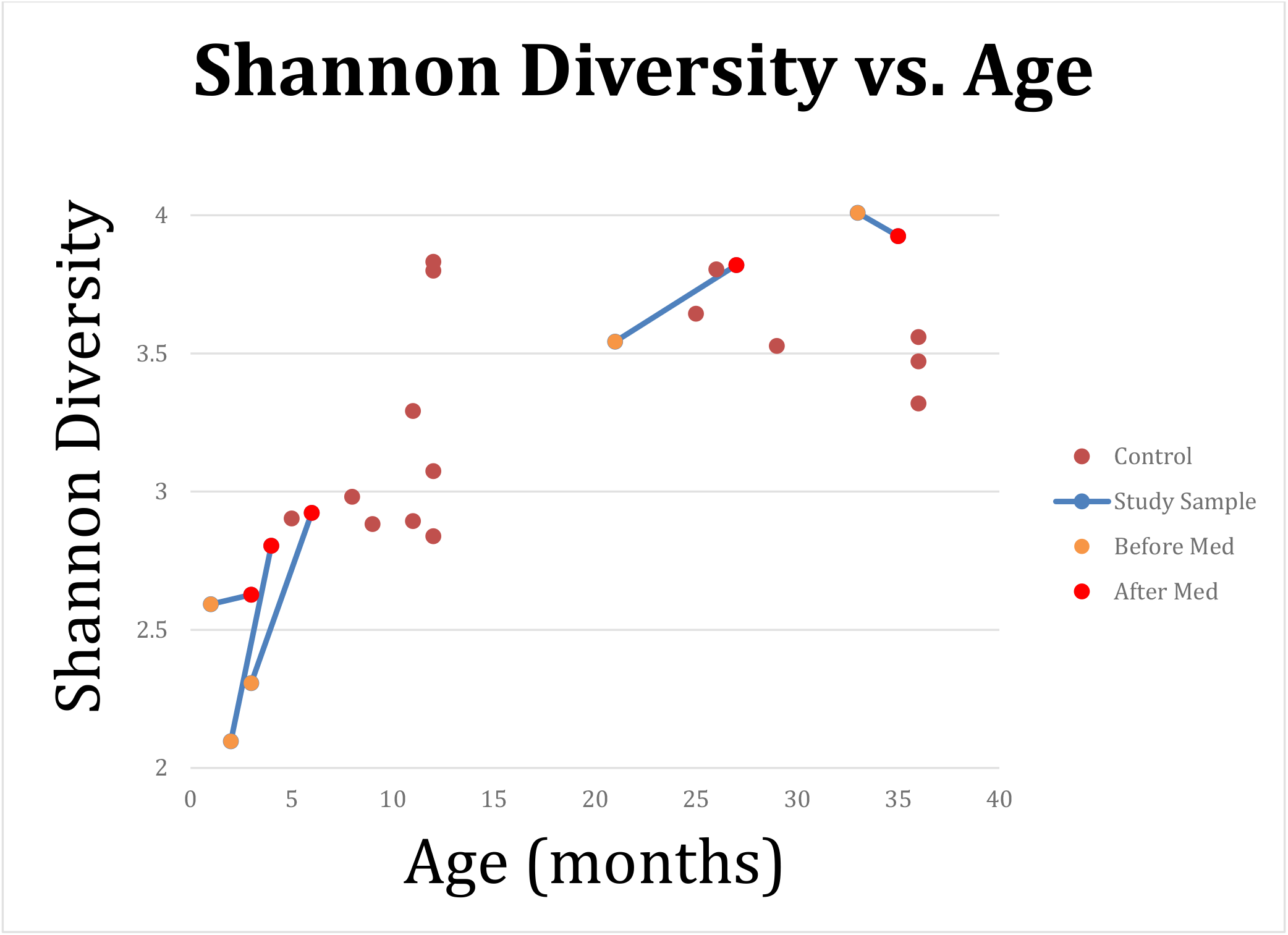
Shannon Diversity of controls and subjects vs. age. The blue lines connect the same subject before and after medication.

#### Beta diversity

We performed PCoA Unifrac weighted analysis on the entirety of the samples (Figure 5). This included two samples from each of the five subjects and three controls (one sample each) for each subject, for a total of 25 samples. Overall, this shows clustering by age. The cluster of samples in the lower left corner represent ages 8 months and greater, whereas the samples in the periphery represent ages 7 months or less (*p* = 0.001).

**Figure 5:**
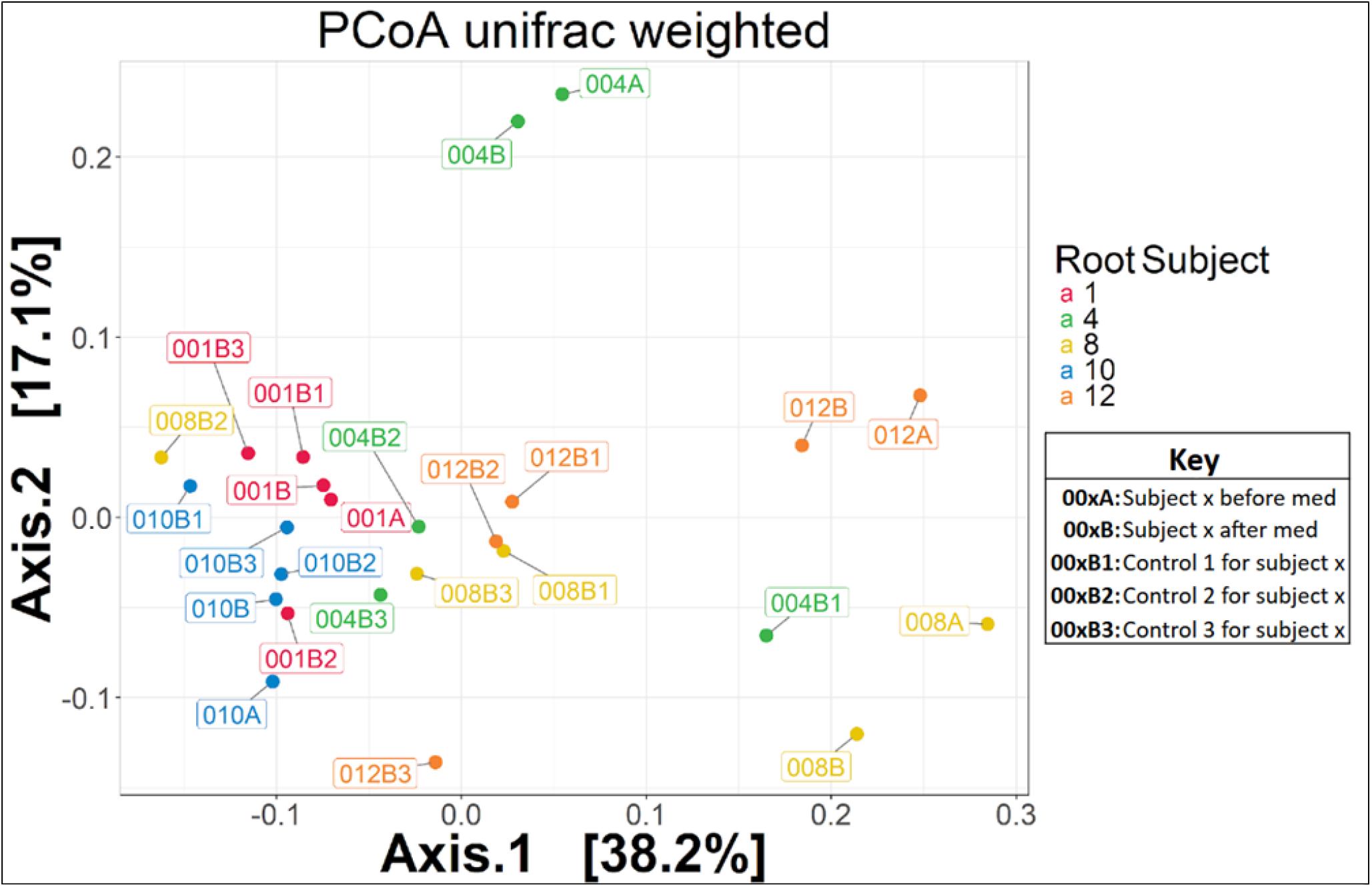
Ordination of experimental and control samples. The data points are color-coded by subject. For example, 001A represents subject 1 before medication. 001B represents subject 1 after medication. 001B1, 001B2, 001B3 represent the three controls for subject 1.

#### Relative abundance of taxa

As part of the control analysis, we compared bacterial abundance of the controls to the post-treatment subjects (Table 5). Among others, we found a lower abundance in the order *Pasteurellales* (*p* = 0.008), the family Lachnospiraceae (*p* = 0.046), and in the genus *Haemophilus* (*p* = 0.001) in post-treatment group.

**Table 5:**
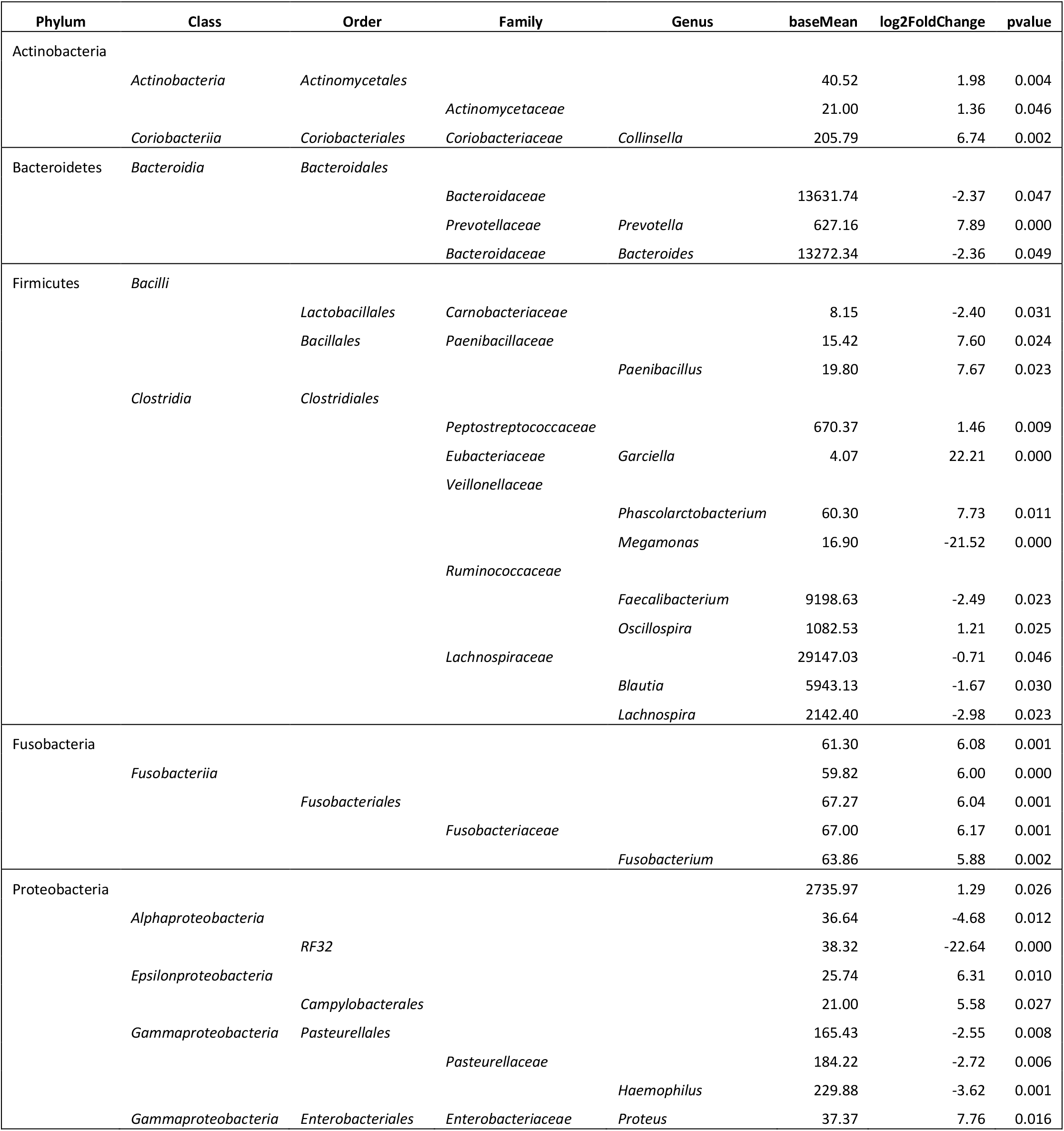
Significant changes in differential abundances between controls and subjects post-treatment. Any taxon with a p-value represents a significant difference in abundance. A negative log2 fold change indicates a lower abundance in subjects post-treatment and a positive log2 fold change indicates a higher abundance in subjects post-treatment.

## Discussion

To the best of our knowledge, we are the first to study the effects of gastric acid suppression on the microbiome of children with the use of age-matched controls, as well as to use qPCR to detect toxigenic and nontoxigenic *C. difficile*. We found a significant increase of *Lactobacillus*, however, we did not detect toxigenic *C. difficile* in our samples. In our control analysis, we found that both alpha and beta diversity were more strongly correlated with age than other factors (presence of GERD, treatment with gastric acid suppression, type of medication).

In 2015 there were two large studies comparing the microbiomes of adults who use PPIs to those who do not. Jackson *et al*. did not find significant changes in alpha diversity when correcting for covariates associated with PPI use such as age, BMI, and antibiotic use (21). Imhann *et al*. did find a significant decrease in alpha diversity with PPI use (22), however did not mention any correction of covariates. There have since been two smaller prospective studies comparing the gut microbiomes of adults before and after PPI use (23,24). While Freedberg *et al*. showed no difference in alpha diversity after PPI use, there were changes in taxa associated with CDI (increased Enterococcaceae and Streptococcaceae, decreased Clostridiales) (23). Hojo *et al*. used quantitative RT-PCR for 22 different taxa. They found significant increases in *Lactobacillus* and Enterobacteriaceae (shown to be associated with CDI in adults (28)), as well as *Streptococcus* and *Staphylococcus*. Of the taxa mentioned in these two studies, our study only replicated the significant increase in *Lactobacillus*.

The only other study investigating the effects of gastric acid suppression on children was in 2017 by Castellani *et al*. They compared the microbiome of infants at three time points: before starting a PPI, 4 weeks into PPI treatment, and 4 weeks after discontinuation of PPI treatment. When comparing baseline microbiome to 4 weeks of PPI treatment, this study showed no significant change in alpha or beta diversity. They found a significant change in alpha diversity when comparing their post-PPI group to both the pre-PPI and during-PPI group (25). Commentary on this study by Drall *et al*. in 2018 suggests that this initial lack of change in diversity may actually represent a deviation from the normal increase in diversity that would occur during microbiome development at this age (29). We also did not observe a significant change in alpha diversity when pooling our subjects together, however, four out of five of our subjects showed an increase in alpha diversity after two months of treatment, following a similar trajectory to their age-matched controls (Figure 4).

Castellani *et al*. compared microbial composition before treatment to 4 weeks into treatment and found a significant decrease in *Lactobacillus* and *Stenotrophomonas* and a significant increase of *Haemophilus*. In contrast, we showed an increase in *Lactobacillus*, as previously shown in adult studies (24), which has also been shown to be positively correlated with CDI in adults (28).

Given our use of age-matched controls, we were able to perform novel analyses. We found that the beta-diversity of our older subjects (> 7 months old) was similar to their age-matched controls (Figure 5). When comparing the taxa of our controls to the post-treatment subjects, we found significantly lower abundances of *Pasteurellales* and *Haemophilus* in the post-treatment group. This was also found when comparing our subjects pre-treatment and post-treatment. This finding supports the notion that this change in microbial abundance is associated more with gastric acid suppression rather than natural progression of the microbiome. We also found a significantly lower abundance of Lachnospiraceae in subjects-post-treatment, which has been shown to be lower in adults with CDI (28). We also performed

*C. difficile* toxin PCR to detect toxigenic *C. difficile*. Nontoxigenic *C. difficile* was present only in one subject both before and after PPI treatment. Surprisingly, no toxigenic *C. difficile* was found even though asymptomatic carriage has a high prevalence at this age (30).

### Limitations

A major limitation is our sample size. The small sample size is due to a decrease in the use of PPIs and H2RAs just as the study was started. With a larger sample of both subjects and controls, we would be able to better characterize normal microbiome development and potential deviations in this development in our experimental group. It is possible that, like Castellani’s study, our lack of change in alpha diversity with PPI use actually represents a deviation from the normal increase in diversity you would see in this age group. In addition, a larger sample size would allow us to better assess whether there are differential microbiome changes between different classes of medication (H2RA vs. PPI).

## Conclusion

The goal of this study was to further characterize the effect of gastric acid suppression medications on the developing microbiome of children. This is a difficult task given the fact that there is variability in the development of the microbiome in the first few years of life, with effects from both intrinsic and extrinsic factors, and no universally agreed-upon “normal” development. With this in mind, we compared our subjects to healthy age-matched controls and found that both the alpha and beta diversities in our subjects resembled the controls, even after use of gastric acid suppression. While we did not detect toxigenic *C. difficile* in our samples, we did find a significant increase of *Lactobacillus*, which has been previously associated with CDI in adults. These pilot results warrant further exploration in a larger cohort.

## Data Availability

All data produced in the present study are available upon reasonable request to the authors

